# Assessment of a 60-biomarker health surveillance panel (HSP) on whole blood from remote sampling devices by targeted LC/MRM-MS and discovery DIA-MS analysis

**DOI:** 10.1101/2023.02.04.23285394

**Authors:** Stephen A. Whelan, Nathan Hendricks, Zachary L. Dwight, Qin Fu, Annie Moradian, Jennifer E. Van Eyk, Susan M. Mockus

## Abstract

**Summary:** Telehealth, accessing healthcare and wellness remotely, should be a cost effective and efficient way for individuals to receive care. The convenience of having a robust remote collection device for blood tests will facilitate access to precision medicine and healthcare. Herein, we tested a 60-biomarker health surveillance panel (HSP), containing 35 FDA/LDT assays and covering at least 14 pathological states, on 8 healthy individuals’ ability to collect their own capillary blood from a lancet finger prick and directly compared to the traditional phlebotomist venous blood and plasma collection methods. Samples were quantitatively analyzed by a liquid chromatography-multiple reaction monitoring-mass spectrometry method targeting 114 peptides representing 60 HSP proteins and by a data-independent acquisition mass spectrometry discovery method, resulting in up to 3,811 protein identifications including 122 FDA approved biomarkers. The >90% similarity between the three biofluids indicate that remote collection devices are a viable option for personal blood proteome biosignature stratification and health analysis.

## Introduction

Precision medicine consists of identifying the disease state of an individual so that an effective intervention or treatment can be prescribed to correct or lesson the deleterious effects of their phenotypic ailments.^1^ Precision health is focused on prevention, maintaining and even optimizing health, which should be an effective way to intervene common sedentary and western diet induced lifestyle metabolic diseases including the onset of age-related diseases.^2–7^ More people are prioritizing and monitoring their personal health regime through an array of health strategies involving nutrition, physical fitness, healthy microbiome, wearable technology, and mental health wellbeing.^8–12^ Physicians, scientists, and individuals can become proactive in their self-health management and requires appropriate assays beyond the conventional medically prescribed tests to quantitatively measure disease progression to provide actionable and personalized health insights. Both precision medicine and precision health have benefited from significant advancement of multi-omics technologies that allow millions of quantitative and qualitative tests in genomics, epigenetics, transcriptomics, proteomics, metabolomics along with the development of powerful computational bioinformatic tools and machine learning algorithms to help understand an individual’s unique phenotype so that more effective interventions may be prescribed, customized and monitored for desired outcomes.^13–17^

The blood proteome is an informative source of biomarkers representing personalized signatures of physiological phenotype, by interacting and responding to the entire body’s cellular tissue disease or health state.^18^ The proteome also represents an individual’s genetic predisposition, continually responds to environmental factors and infectious agents as well as drug treatments, physical exercise^9,10,19,20^ and nutritional intervention.^11,12,21^ In addition, the proteome’s unique isoforms, post-translational modifications (e.g. A1C),^22^ longer half-lives compared to metabolites, and shorter half-lives responsive to acute stimulus (e.g. CRP)^23^ allows for specific and stable targeted diagnostic tests to be developed.^22,23^ There has been significant development in biomarker assays targeting the proteome for health or disease by companies such as Olink^24,25^, MRM Proteomics,^26^ Biognosys,^27^ Somalogic^28^ and Seer.^29^ Similarly, at Cedars-Sinai Medical Center, the Advanced Clinical Biosystems Research Institute (ACBRI) and Precision Biomarker Laboratories (PBL) have developed a 60 protein health surveillance panel (HSP) that includes 35 FDA and LDT protein clinical assays^30^ and covers 14 physiological diseases. The HSP multiplex is composed of 1-2 peptide per protein with a quantifier (used to calculate protein concentration) and when possible, a qualifier peptide (confirms protein result).^31^ The current data collected via liquid chromatography-multiple reaction monitoring mass spectrometry (LC-MRM/MS)^32^ and discovery data-independent acquisition mass spectrometry (DIA-MS)^33^ analysis with stable isotopically labelled (SIL) peptides, is encouraging for the future expansion to larger multiplex biomarker panels into the hundreds of proteins to provide a more comprehensive health and disease assessment than is currently available with current standard-of-care medicine.^34^

Most health assessment screens require multiple blood draws collected at a healthcare facility by a trained phlebotomist. Samples are then, packed for transport and delivered to a laboratory service provider for sample processing and analysis. The benefits of having access to a safe, viable and reliable remote collection device is convenient and cost effective for patients, physicians and clinics, allowing for an array of blood tests.^35–37^ Biomarker analysis has primarily been assessed with serum or plasma, where all cellular mass including red blood cells, white blood cells, platelets are removed to reduce the complexity of the biofluid.^18^ We found that the Mitra device (Neoteryx) which absorbs 10µl^38^ when collected correctly through its volumetric absorptive microsampling (VAMS) technology,^39,40^ facilitates self-administered blood collection from a finger lancet needle prick with highly reproducible replicate collection,^35,41^ as demonstrated with very low coefficients of variation (%CV). The Mitra devices have been shown to be successfully utilized with many conventional diagnostic tests currently ordered by a physician and can be easily adapted for automation.^42,43^ The stability of whole blood on Mitra devices on a subset of proteins has also been assessed by multiple groups, including Van Eyk *et al*..^43–45^ The Mitra device allows testing of infectious diseases,^46^ small molecule drugs,^47^ therapeutics,^48,49^ remote patient monitoring,^50^ pediatric pharmacokinetic studies,^49^ hormones, and newborns to name a few key areas of health. In addition, an inexpensive, easily self-assisted remote collection device will allow people in a health care system as well as people in underserved populations and remote regions of the world to have access to quality medical care, clinical grade assays and treatment monitoring.^40,44,51,52^

In this study, we show that the HSP multiplex assay quantitative results between capillary blood, venous blood, and plasma indicate the use of remote collection devices is a viable option for health analysis and that each individual person has a unique proteome biosignature. The convenience to patients, array of circulating tests and sample processing efficiency on a Mitra device in addition to the technological advancements in mass spectrometry and bioinformatic tools to quantitively identify thousands of physiological relevant proteins in whole blood may shift the paradigm away from analyzing plasma alone.

## Results

### Sample collection and LC/MRM-MS targeted analysis

We tested an internally developed 60 protein HSP multiplex assay on 8 volunteers consisting of 5 males and 3 females ages 27 to 60. Volunteers were required to collect their own capillary whole blood from a lancet finger prick using Mitra devices which was compared to the traditional phlebotomy method of venous whole blood collection and plasma isolation, all samples collected on the same day. All samples from the three biofluids were analyzed by both targeted LC/MRM-MS and DIA analysis (Figure 1A and 1B). Each Volunteer collected six Mitra devices of a capillary blood drop (10µl) from a lancet finger prick that was matched to 10µl of venous whole blood pipetted onto six Mitra devices on the same day. According to the American Red Cross, blood is estimated to be 55% plasma, we used 5.5 µl of matched isolated plasma for in-solution digestion for a theoretical 1:1:1 (capillary:venous:plasma) comparison (Figure 1A). A buffer of 33% TFE in 100mM ammonium bicarbonate was determined internally to yield the best results. Briefly, Mitra tips were removed and placed into 96 well plates (Figure 1A and Figure S1), trypsin digested, and a stock containing 40 fmol of each Stable Isotopic Labelled (SIL, Table S1), representing each targeted peptide was spiked into solution for every 1 µg of plasma protein. The equivalent of 7.5 µg of plasma protein and 300 fmol of SIL was loaded onto the column and analyzed by targeted LC/MRM-MS using a 3000 Ultimate HPLC interfaced with a Sciex 6500+ triple quadrupole mass spectrometer (Figure 1B). MS raw data was normalized and searched by using our in-house ProEpic™ software platform, with smart peak^53^ used to identify the quality of the peaks. ProEpic™ software calculated peak area ratios (PAR = endogenous peptide/SIL) for all 114 peptides representing 60 HSP biomarker proteins across all 8 volunteers capillary blood (each volunteer n=6, total 48 samples), venous blood (each volunteer n=6, total 48 samples) and plasma samples (each volunteer n=3, total 24 samples). The %CV ((standard deviation / mean) x 100) was calculated from across each volunteer’s 6 replicates for the quantifier peptides (Table S2). For simplicity, we compared the biological replicates averages of the quantifier peptide transitions of all HSP proteins between the capillary blood, venous blood and matched plasma and all were below 20%CV (Figure 2A and Table S2). The average %CV for the quantifier peptides for capillary blood was 10.92%, for venous blood was 5.92%, and plasma was 5.76% (Table S2). There were four sets of quality control (QC) groups, pooled from the digestion of capillary blood of all 8 volunteers biological replicates (n=48), pooled from the digestion of venous blood of all 8 volunteers biological replicates (n=48), pooled from the digestion of plasma of all 8 volunteers (isolated from venous blood) biological replicates (n=24), and the purchased human pooled plasma (pool4) used at the start, middle and end of each sample plate analysis had an overall average coefficient variance (or relative standard deviation (RSD)) of 8.82 for pool4 plasma, 8.68 for volunteer pooled plasma, of 8.64 for pooled capillary blood from Mitra devices and 9.12 for pooled venous blood from Mitra devices (Figure S2, raw data in Table S3).

**Figure 1.**
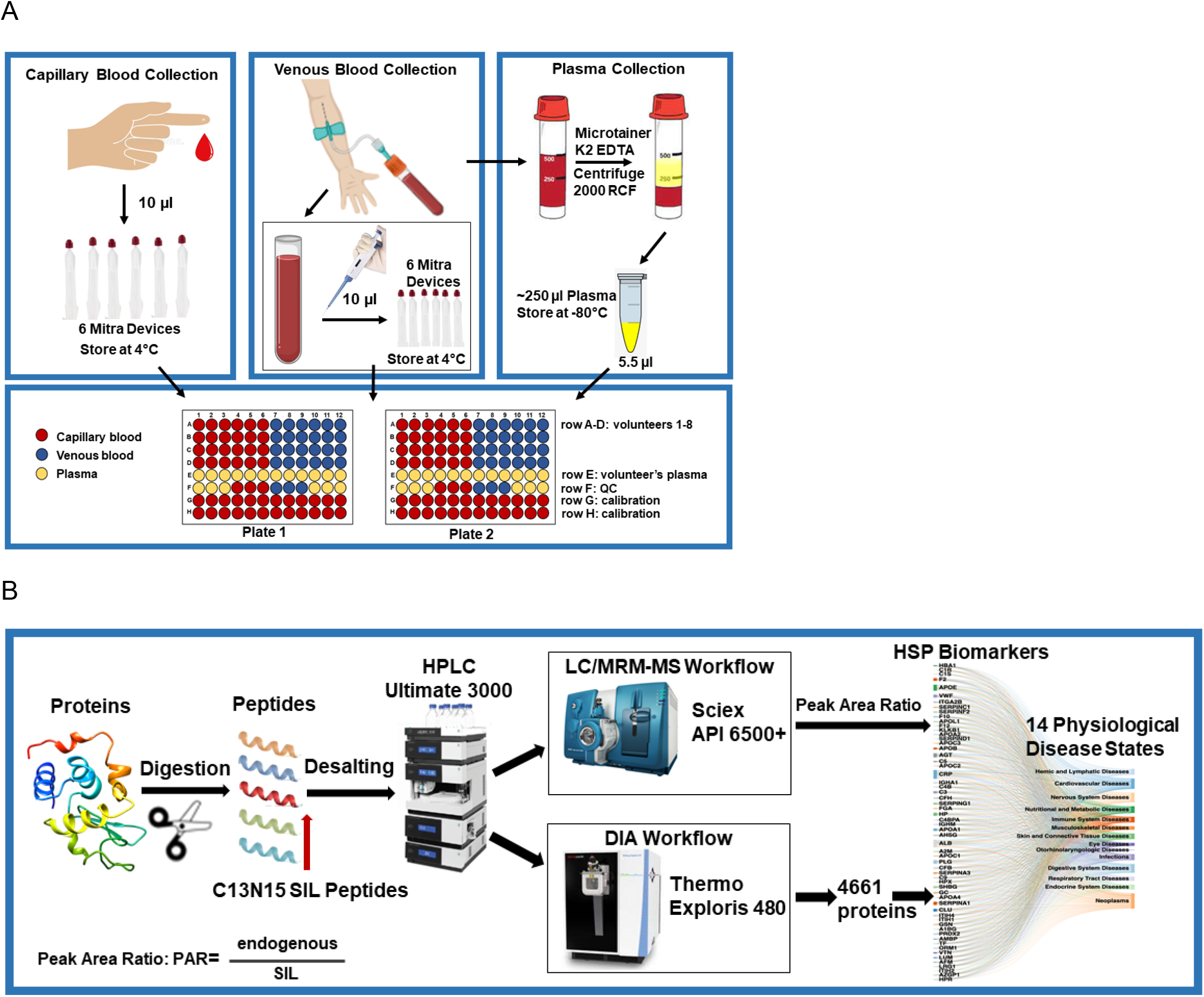
Health Surveillance Panel (HSP) workflow overview. **(A)** Outline of capillary blood (n=6) and venous blood (n=6) Mitra Device collection matched to plasma (n=3) from 8 volunteers. (**B)** Outline of LC/MRM-MS platform consisting of the Thermo Ultimate 3000 HPLC and Sciex 6500+ triple quadrupole mass spectrometer and DIA discovery platform consistening of the Thermo Ultimate 3000 and Exploris 480 mass spectrometer.

**Figure 2.**
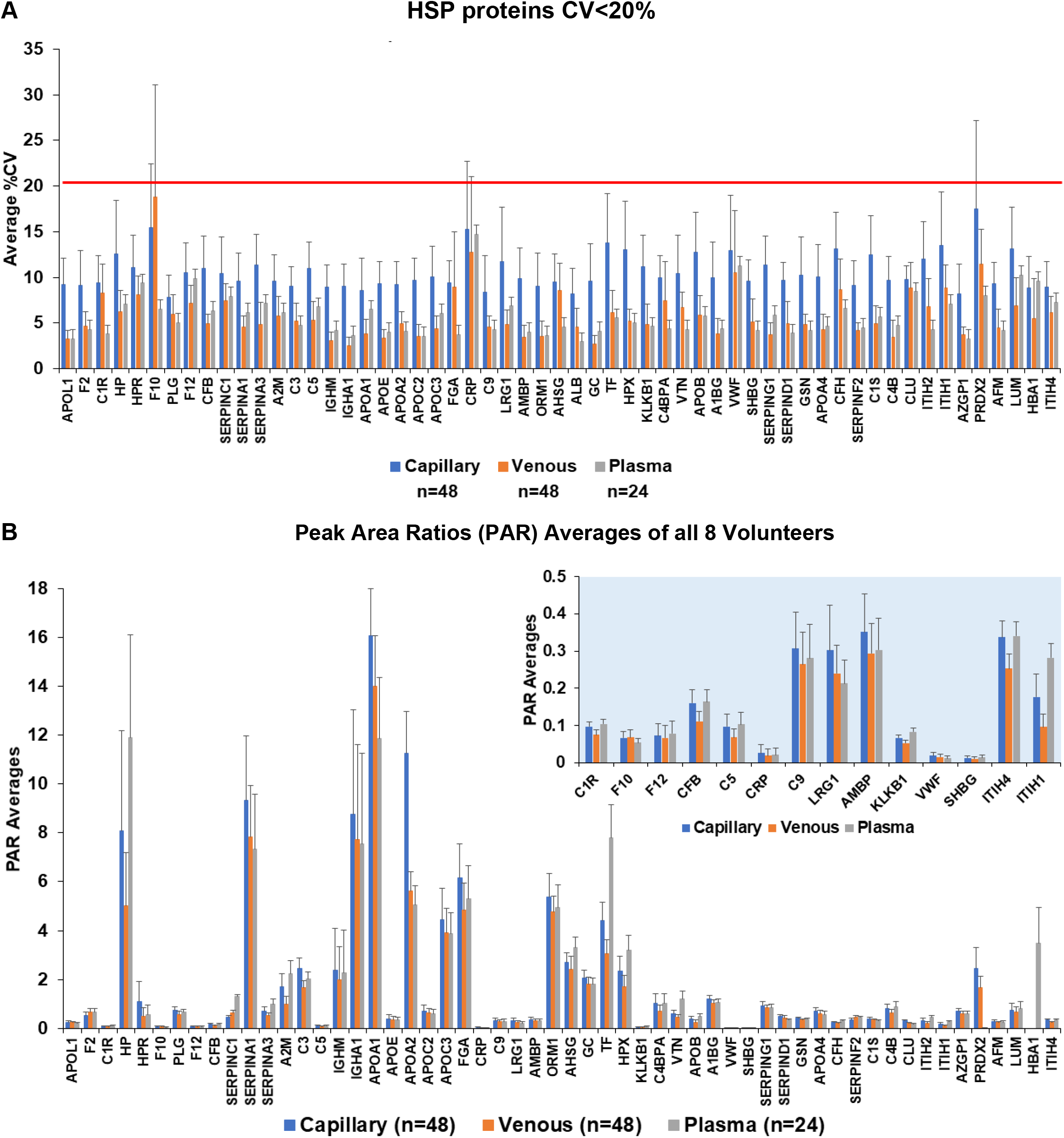
Overall %CV and PAR averages of 8 volunteers HSP proteins in capillary blood, venous blood and plasma. **(A)** The average of all 8 volunteers six biological replicates of capillary blood (n=48), six replicates of venous blood (n=48) and 3 replicates of matching plasma (n=24) for each biomarker in the HSP was <20%CV in all three biofluids. (**B)** Comparison of PAR averages from 8 volunteers and replicates for each HSP protein from whole blood capillary (n=48), whole blood venous (n=48) and matched plasma (n=24).

The average PAR of all 8 volunteers biological replicates were compared to determine if there were any major differences between the three biofluid collection methods (Figure 2B, raw data in Table S2). The majority of HSP proteins demonstrated a relatively similar PAR intensity between the biofluids. Albumin (ALB P02768) was not plotted since it is the most abundant plasma protein and compared to the other HSP proteins would be off scale. In addition, hemoglobin (HBA P69905) and peroxiredoxin 2 (PRDX2 P32119) from capillary and venous blood was excluded since it is found in very high abundance in red blood cells and would also be off scale. However, hemoglobin from plasma was included to demonstrate the relative quantity compared to other HSP proteins (Figure 2B). The lower abundant HSP proteins were highlighted in a zoomed graph so that the relative quantities could be seen between biofluids (Figure 2B). Interestingly, each volunteer had a unique PAR intensity biosignature pattern for many of the HSP proteins (e.g. CRP, VWF, SHBG, APOE, IGHA1, ITIH1, HP, CLUS and GSN to highlight a few) which was consistent across each biofluid of capillary blood, venous blood and plasma (Figure 3A and 3B). One of the major benefits of collecting whole blood on Mitra devices, is that differential hemolysis due to phlebotomist collection and plasma isolation is avoided for the red blood cell specific proteins HBA and PRDX2 (Figure 3B). This was demonstrated in each volunteer having a similar PAR intensity biosignature for HBA and PRDX2 in both capillary and venous blood. When we looked at the levels of HBA and PRDX2 in plasma for each volunteer the PAR did not match either the capillary or venous pattern, however the HBA pattern matched the PRDX2 PAR pattern for each volunteer in the plasma samples, indicating inadvertent hemolysis typically seen in plasma collection (Figure 3B). This observation demonstrates that even a small amount of red blood cell perturbation results in significant differences in plasma levels related to mechanical fractionation of plasma rather than biological differences. HBA PAR and PRDX2 PAR is upward to 1000x and >200x, respectively more concentrated in red blood cells than plasma, where even if a small amount of hemolysis occurs it greatly affects their concentrations in plasma, demonstrating the added benefit of using whole blood for these two HSP proteins in determining biological relevance.

**Figure 3.**
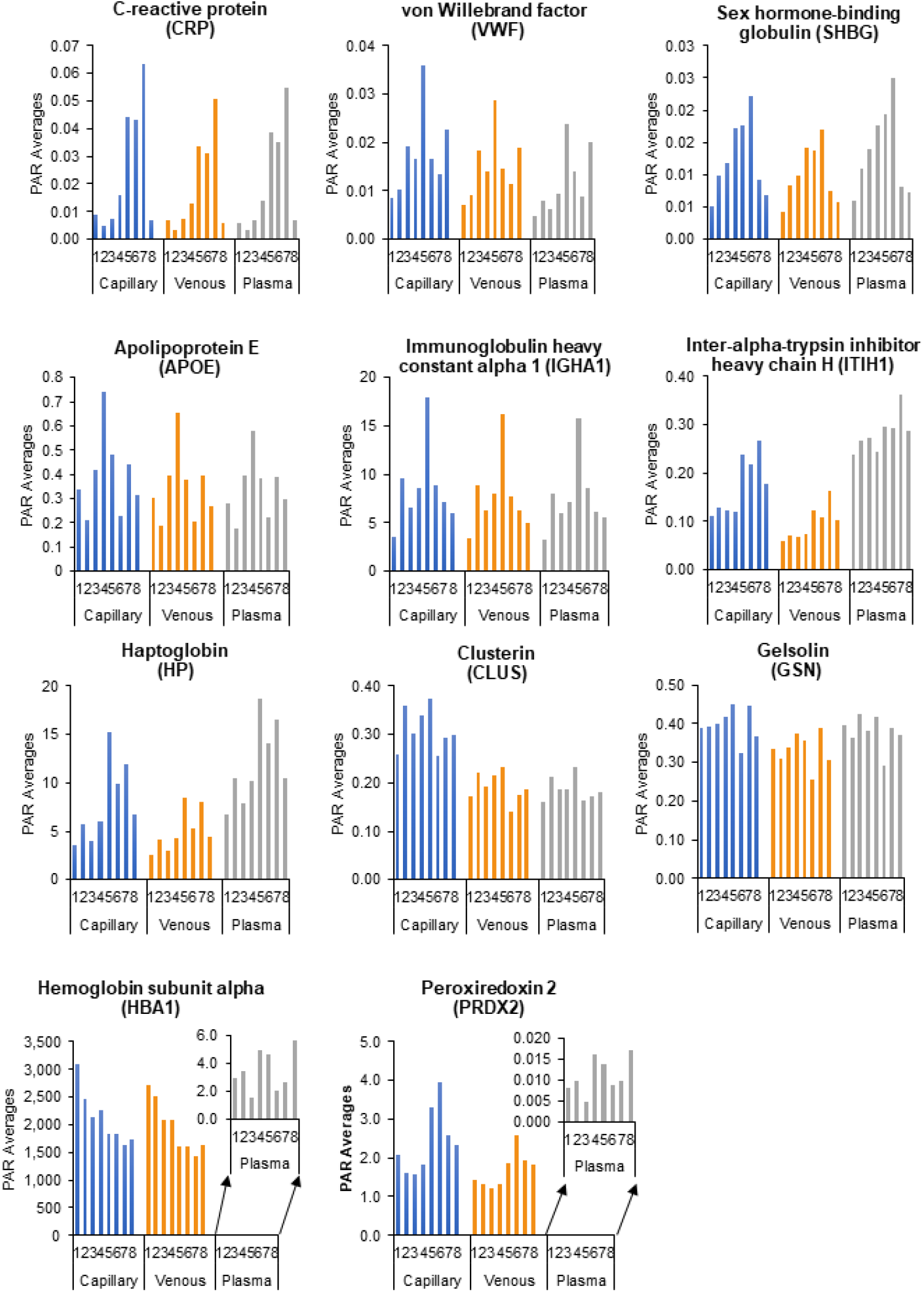
Each volunteer has a unique HSP protein biosignature, that is similar across three biofluids. **(A)** Comparison of average PAR for each volunteer (PAR represents capillary n=6, venous n=6, plasma n=3) for C-reactive protein (CRP), von Willebrand factor (VWF), sex-hormone binding globulin (SHBG), apolipoprotein E (APOE), immunoglobin heavy constant alpha 1 (IGHA1), inter-alpha- trypsin inhibitor heavy chain H (ITIH1), haptoglobin (HP), clusterin (CLUS), and gelsolin (GSN). **(B)** The red blood cell proteins hemoglobin (HBA) and peroxiredoxin 2 (PRDX2) are significantly higher abundant in whole blood collected from capillary blood and venous blood, which share the same biosignature intensities. However, in plasma both HBA and PRDX2 are far less abundant, and the volunteers do not share the same biosignature intensity as capillary blood and venous blood, which is likely due to minor differences in plasma isolation as can be seen with HBA and PRDX2 having similar profiles in only the plasma biofluid.

### Statistical analysis of HSP proteins similarity in capillary blood, venous blood and plasma

The PAR data of all 8 volunteers including all their biological replicates were analyzed by several statistical techniques to determine if there was any inconsistent variance from the other volunteers when compared to the different biofluid collection methods. In Figure S4, the Pearson correlation coefficient was plotted against p-values for all the HSP proteins to demonstrate the similarity of most HSP proteins in each biofluid comparison and highlights the proteins that are significantly different. Only three proteins, (SERPINC1, SERPINF2, and C1R) did not have a p-value ≤0.05 when comparing capillary whole blood to venous whole blood collected on Mitra devices (Figure S3A and Table S4). As expected, HBA and PRDX2 were similarly variable in both the comparisons of capillary and venous whole blood to plasma due to their known high abundance in red blood cells (Figure S3B and S3C). In addition, more differences were seen in the comparison of whole blood to plasma (SERPIND1, APOA1, APOA2, FGA, and ALB, Table S4).

### Protein biosignatures of 8 healthy/normal individuals

One benefit of having a 60 protein biomarker HSP multiplex assay is the ability to correlate specific changes in the HSP protein intensities to distinguish unique biosignature profiles in normal healthy individuals, similar to the 8 volunteer cohort studied here. The HSP protein data was used to analyze each volunteer’s set of biological replicates within each biofluid of capillary blood (n=6), venous blood (n=6) and matched plasma (n=3) in comparison to all the volunteers. The heatmap analysis demonstrated that each individual volunteer had a unique biosignature of upregulated and downregulated proteins in each biofluid (Figure 4). Three of the volunteers (5, 6 and 7) had a number of inflammatory proteins including CRP upregulated. Interestingly, volunteer 5’s results correlated with a same day blood analysis of Covid 19 IgG antibodies and N capsid antibodies that indicated a past infection (Table S5). The other two volunteers also had a history of mild inflammation and hypertension.

**Figure 4.**
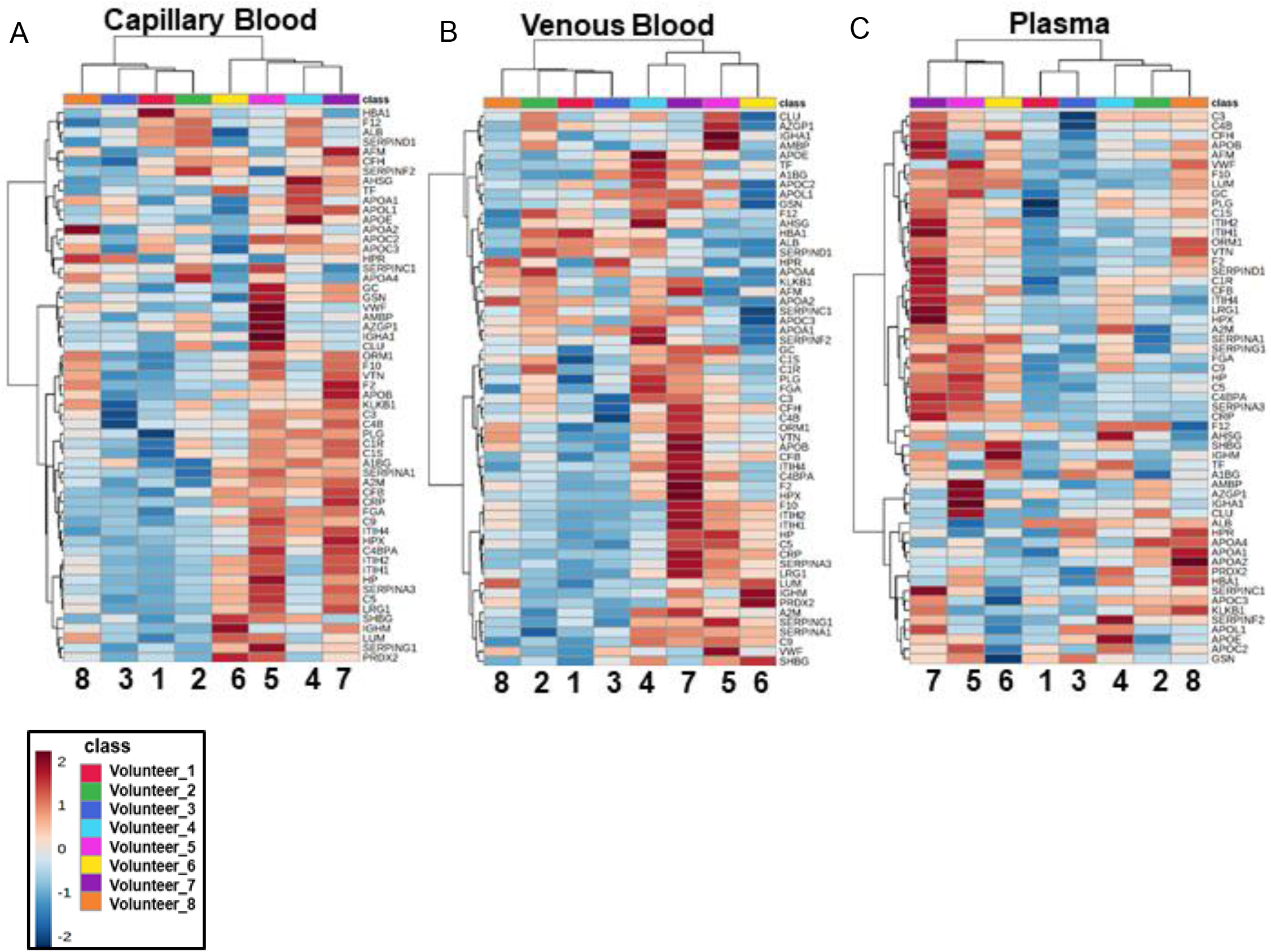
Each volunteer has a unique biomarker biosignature across capillary blood, venous blood and plasma. Hierarchical clustering heatmaps distribution of HSP proteins from 8 volunteers **(A)** capillary whole blood, **(B)** venous whole blood and **(C)** plasma biofluid. The averages of replicates were grouped for each volunteer using the Euclidean distance measure and Ward clustering from T- test/NOVA calculations.

Since we did not have an established disease cohort to compare within this study we further tested if replicates from capillary blood collected on Mitra devices (n=6) would cluster with replicates from venous blood (n=6) collected on Mitra devices (Figure 5). Volunteer 5, capillary and venous blood replicates all clustered together in Cluster 1 distinctly away from the other volunteers (Figure 5). In addition, volunteer 6 and volunteer 7, that have respectively reported a history of hypertension and inflammation, biological replicates from their capillary blood and venous blood both clustered and overlapped in Cluster 2 and Cluster 6. While an active healthy male volunteer 2’s biological replicates clustered in cluster 8, and two other healthy male volunteer 1 in Cluster 4 and volunteer 8 in Cluster 7 overlapped, as did two healthy female volunteer 3 and volunteer 4 clustered near each other encompassed by the red dotted line. Dendrogram hierarchical clustering of all volunteer’s biological replicates demonstrated the replicates clustered for each volunteer in both their capillary blood and venous blood (Figure S4).

**Figure 5.**
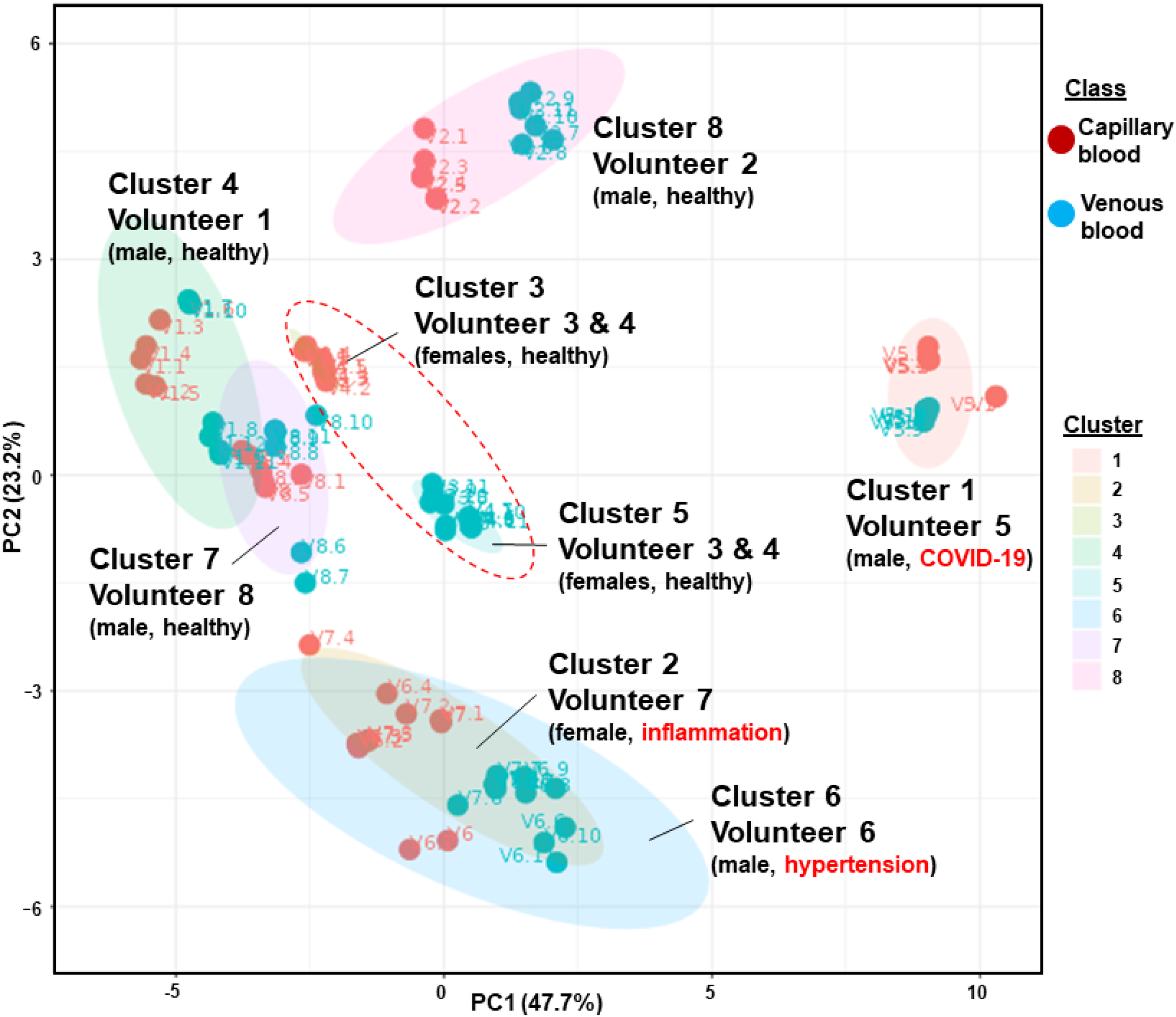
Distinct volunteer biosignature clustering of capillary blood and venous blood. A) K- means clustering of the 8 volunteers biological replicate samples from both capillary blood (n=6) and venous blood (n=6). Gender and general health status listed with each volunteer. Volunteer 5 capillary blood and venous blood biological replicates clustered together and significantly different to the other volunteers. Volunteer 5 had a blood draw for the EMBARC study the day samples were collected for this study and antibody results suggested a past covid 19 infection (see Table S). Volunteers 6 and volunteer 7 reported a history of hypertension and inflammation, respectively, and their biological samples overlapped in Cluster 2 and Cluster 6. The red dotted circle encompassing Cluster 3 and Cluster 5 are two healthy females where their capillary blood clustered and venous blood clustered. Two healthy males volunteer 1 in Cluster 4 and volunteer 8 in Cluster 7 clustered similarly.

### HSP physiological concentration in capillary blood, venous blood and plasma

To determine if the volunteers physiological endogenous HSP proteins were within the linear range of calibration, SIL peptides were spiked in duplicate into pooled tryptic digested capillary whole blood from all 8 volunteers to create calibration curves on both sample plate 1 and plate 2. The sample plates were analyzed on two harmonized Sciex 6500+ triple quadrupole mass spectrometers. Linear regression was used to calculate the slopes for each transition and analyzed in duplicate on each plate. The four calculated slopes for each transition were averaged and the %CV calculated to determine the reproducibility between the calibration curves on two different sample plates analyzed, resulting in 53 of the averaged slopes having <20%CV (Table S6). The SIL spiked into each sample results in 300 fmol analyzed on column, which is in the linear range of almost all HSP proteins. Since the 300 fmol of SIL is in the linear range of the calibration curve for *most* HSP proteins, the PAR for each HSP protein that is also in the linear range of the calibration curve can be used to calculate the endogenous concentration, by multiplying each HSP proteins PAR (peptide/SIL) by 300 fmol (Table S7). The endogenous protein concentration in plasma was calculated from the average PAR from all 8 volunteers and their six replicates (n=48) for each HSP protein and compared to published physiological levels of each protein from the Plasma Proteome Database (http://www.plasmaproteomedatabase.org).^54^ Interestingly, the majority of the volunteers’ HSP proteins were within published physiological ranges^55,56^ and listed in the Plasma Proteome Database.

In addition, the CRP (mg/L) concentrations for each volunteer’s biological replicates in capillary, venous and plasma was calculated and plotted in a box and whisker graph to compare the detected levels within each biofluid (Figure 6A). Interestingly, the three volunteers, 5, 6, and 7 all had elevated physiological levels approaching 10mg/L indicating inflammation. This CRP elevated response correlated with perturbances in inflammatory proteins. All other healthy volunteers had normal CRP levels (about 0.5-1mg/mL). The elevation in CRP demonstrates its utility as a biomarker within the Health Surveillance Panel for detecting inflammatory responses in three of the volunteers within a normal healthy cohort. Since ALB is the most abundant protein in blood and accounts for about 50% of plasma proteins we calculated the concentration for all 8 volunteers and corresponding replicates present in capillary blood (n=48), venous blood (n=48) and plasma (n=24) and compared the three biofluids in a box whisker graph (Figure 6B). In addition, the average of all HSP proteins were calculated from the capillary blood (blue), venous blood (orange) and plasma (grey) samples of all 8 volunteers and corresponding six replicates and were plotted (except the very high abundant HBA and ALB) so that the dynamic range of HSP concentrations could be visualized (Figure 6C).

**Figure 6.**
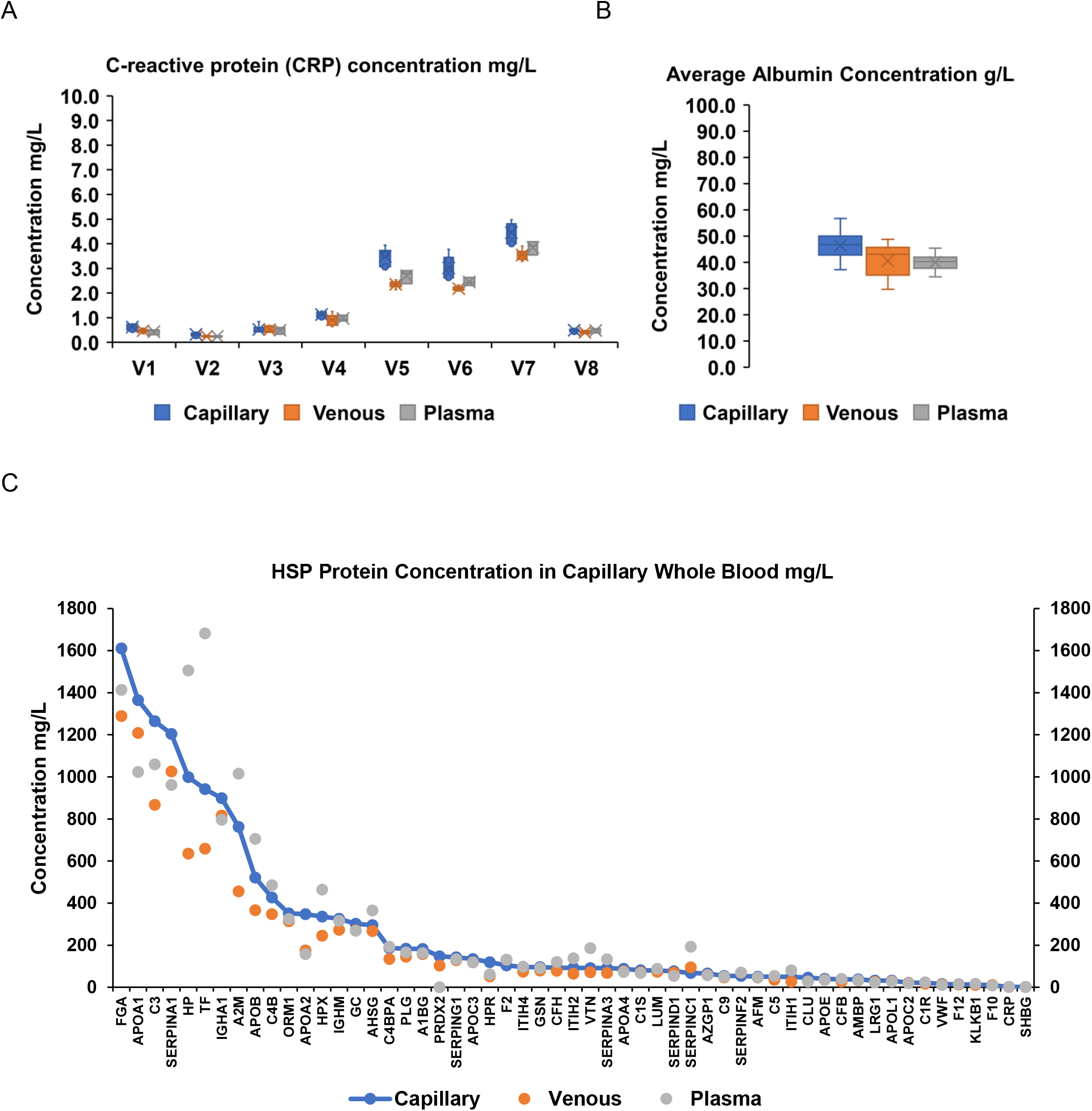
Comparison of physiological concentrations of HSP proteins in capillary blood, venous blood and plasma. **(A)** Comparison of the concentration of CRP (mg/L) in the 8 volunteers capillary blood (n=6), venous blood (n=6) and plasma (n=3). **(B)** Whisker and box plot of the concentration of albumin in all 8 volunteers replicates for capillary blood (n=48), venous blood (n=48) and plasma (n=24). **(C)** Average concentration of HSP proteins of all 8 volunteers replicates in capillary blood (n=48, blue), venous blood (n=48, orange) and plasma (n=24, gray).

### Data-independent analysis (DIA) of capillary blood, venous blood and plasma

Duplicate sample plates were analyzed via the DIA-MS platform using the Exploris 480 mass spectrometer (Figure 1B) to determine if the HSP proteins could also be identified in an unbiased manner in whole blood samples. Using the twin plasma spectral library^57^ and setting a criterion for proteins to exist in >66% of the samples, 1,121 total unambiguous proteins were identified, 396 proteins were observed in capillary blood, 399 were observed in venous blood, and 388 proteins were observed in plasma (Figure 7A, raw data in Table S8). Using the pan-human spectral library^58^ which incorporated many of the proteins found in whole blood cells, a total of 3,811 unambiguous proteins at 1%FDR (Table S9) were identified (4661 proteins were identified when FDR was set to 5%, Table S9), while setting a criteria for proteins to exist in >66% of the samples, 877 proteins were observed in the capillary blood, 1029 in the venous blood and 364 in the plasma (Figure 7A). There are significantly more proteins identified in all three biofluids using the pan-human spectral library. The DIA-MS analysis detected almost all HSP proteins in all three biofluids, except CRP and ITA2B (ITA2B also not detected in the MRM assay, abundance too low), while SHBG and F10 were not seen in both capillary blood and venous blood. In addition, many other biologically relevant proteins were also identified in the whole blood, presumably from red blood cells, white blood cells, platelets, etc. that are typically too low abundant in plasma alone. The DIA identified proteins in capillary blood, venous blood and plasma were compared and ranked in abundance (Table S10).

**Figure 7.**
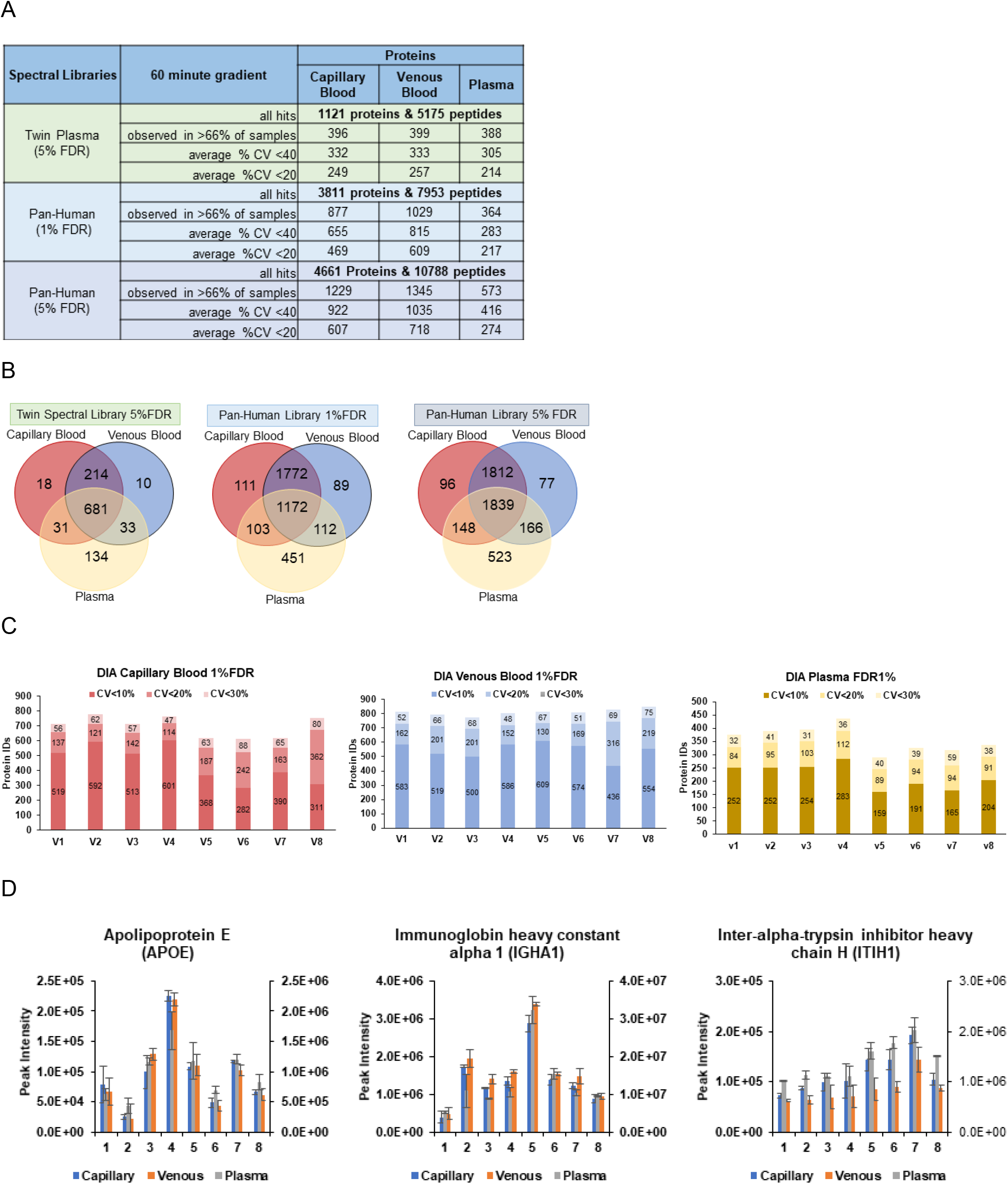
Data independent acquisition (DIA) of all 8 volunteers capillary blood, venous blood and plasma. Data was collected on an Ultimate 3000 and Exploris 480. **(A)** Total proteins identified using both the twin plasma spectral library and pan-human spectral library at 1%FDR and 5%FDR. B) Venn diagram of total proteins identified in capillary blood, venous blood and plasma and Venn diagram of total proteins from twin plasma spectral library search, pan-human spectral library search 1%FDR and pan-human spectral library search 5%FDR. **(C)** Column graph of coefficient of variance of proteins identified by DIA in 3 biological replicates for each volunteer in capillary blood, venous blood and plasma (volunteer 5 in capillary and plasma volunteer 4 is n=2). **(D)** Each volunteer has a unique biosignature across biofluids as seen with the average peak intensities from APOE, IGHA1 and ITIH1 (n=3) which is also a similar pattern as seen in the LC/MRM-MS targeted analysis in Figure 4.

The overlap of the total identified proteins in the three biofluids were compared in Venn diagrams for the twin library and pan-human library (1%FDR and 5%FDR, Figure 7B). The DIA analysis using the pan-human library at 1%FDR resulted in the shared identification of 2944 protein between capillary blood (93%) and venous blood (94%) while plasma alone had 451 (25%) unique proteins identified, indicating although plasma shared 1172 identified proteins with both capillary blood and venous blood, that as expected, plasma and whole blood (which again is ∼55% plasma) had distinct proteomes due to the cellular differences of each biofluid. By increasing the DIA search results FDR filter to 5% (which also increases false positives), plasma shared proteins with capillary and venous blood increased by 661 proteins while only 40 shared proteins increased between capillary blood and venous blood, possibly indicating that there is actually greater overlap between the three biofluids, that get filtered out with higher stringency. Using the data from the pan-human library search with the stringent 1%FDR filter, the identified proteins with CV<10%, CV<20% and CV<30% were compiled into stacked columns for each individual volunteer into three separate charts for capillary blood, venous blood and plasma (Figure 7C) in order to visualize the upper limits of reproducible relative quantitation in each biofluid. The DIA analysis is able to reproducibly quantitate approximately 600-700 proteins in capillary blood, ∼800 proteins in venous blood and ∼300-400 proteins in plasma. The DIA analysis also demonstrated reproducible quantitation of APOE, IGHA1 and ITIH1 (n=3) in each biofluid for each volunteer and each volunteer had a unique biosignature (Figure 7D) similar to that seen in Figure 4 targeted analysis. The DIA-MS data also contained at least 122 FDA approved biomarkers between all three biofluids (Figure 8). The average %CV across all 8 volunteers (3 biological replicates, n=24) was calculated for each FDA approved biomarker and plotted for each biofluid to observe which biomarkers in the DIA-MS analysis had the least variation with <10%CV, <20%CV, and <40%CV. Surprisingly without SILs, the capillary blood, venous blood and plasma, respectively had 30, 38 and 36 FDA biomarkers <20%CV. In the future, choosing the most reliable peptides for each biomarker and creating SILs will decrease the variance. A total of 83 FDA biomarkers were shared between the three biofluids, while 27 and 5 FDA biomarkers were seen in whole blood and plasma, respectively (Figure 8B). A comparison of whether traditional plasma collection or whole blood with a remote collection device is used there is an opportunity to reproducibly quantitate hundreds of proteins by creating targeted assays (with either MRM on triple quadrupole mass spectrometers or parallel reaction monitoring (PRM) on orbitrap mass spectrometers) lists representing 700-800 to well over 1000 proteins with the addition of stable-isotope-labelled (SIL) peptides in order to obtain comprehensive proteomic biosignature assays.

**Figure 8.**
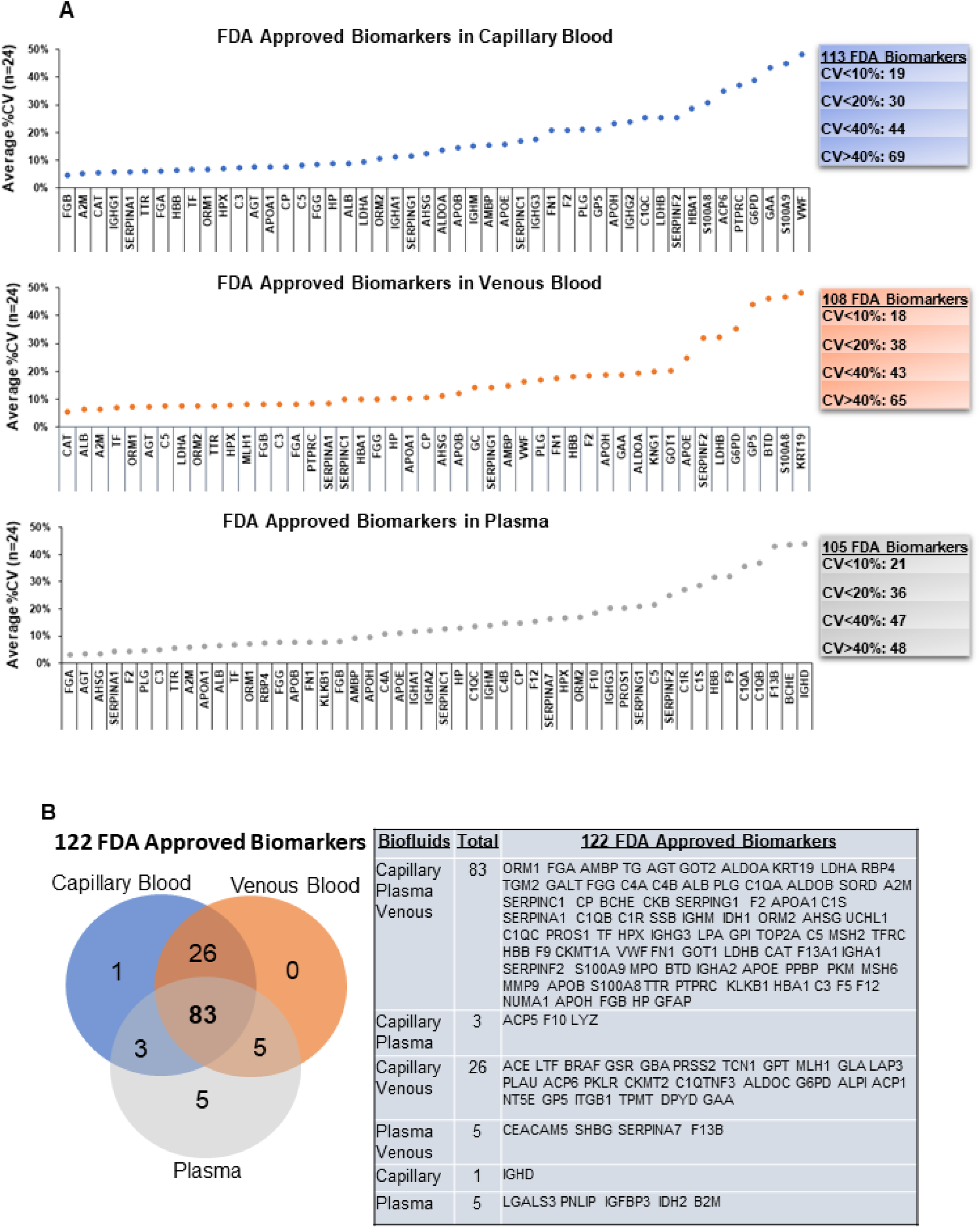
At least 122 FDA approved biomarkers are identified in the DIA-MS analysis of the three biofluids. **A)** Capillary blood contained 113 FDA biomarkers with 19<10%CV, 30<20%CV, 44<40%CV and 69>40%CV. Venous blood contained 108 FDA biomarkers with 18<10%CV, 38<20%CV, 43<40%, and 65<40%CV. Plasma contained 105 FDA biomarkers with 21<10%CV, 36<20%CV, 47<40%CV, and 48>40%CV. Each FDA biomarker is the calculated average %CV of all 8 volunteers (n=24) and only the biomarkers with CV’s<50% were plotted. **B)** Venn diagram the FDA approved biomarkers in capillary blood, venous blood and plasma demonstrating 83 shared with all three biofluids, 27 in whole blood and 5 in plasma alone. All FDA biomarkers are listed for each biofluid.

## Discussion

The combination of having a targeted 60 protein HSP multiplex assay and the convenience of a robust remote collection device will facilitate access to precision medicine and precision health. The HSP assay consisting of approximately 35 FDA and LDT protein biomarker clinical assays^30^ (Table S7) and representing 14 physiological diseases is an excellent starting point for covering a broad range of biologically relevant proteins to assess overall health of an individual. We determined that there were quantitative similarities between the three biofluids analyzed with the HSP assay. The ability of an individual to lance their own finger and collect a drop of blood cannot be underestimated. Previous and our preliminary studies indicated not only that people are squeamish about pricking their own finger but also are not able to reliably collect 60-100µl of their own capillary blood required from some remote collection devices such as the Noviplex cards (Telimmune). Never mind the traditional microtainers that require 250µl of blood, which pose even more of a challenge for self-collection. One major benefit of the Mitra device is that its VAMS technology allows for the accurate measurement of 10µl of a single drop of blood which resulted in an overall coefficient of variance for each HSP protein well below 20% CV. Each Volunteer collected six biological replicates which was directly compared to a venous blood draw pipetted onto six Mitra devices and compared to plasma isolated from the same venous blood draw, all within 24 hours to assess variability in the biofluid analysis rather than the collection time. The HSP proteins in all three biofluids had coefficient of variance well below 20%, demonstrating the overall robustness of the HSP targeted assay and that large multiplex biomarker panels are possible with remote collection devices. Conducting a targeted analysis using SIL peptides for each protein corrects for any sample processing from the time of desalting, drying peptides down, resolubilizing and any variability during the instrument analysis or from different LC/MRM-MS system platforms. The four sets of QCs consisting of purchased plasma, pooled volunteer plasma, pooled peptides from capillary blood and pooled peptides from venous blood were analyzed at the beginning, middle and end of each sample plate. Each sample plate was analyzed on two harmonized LC/MRM-MS systems on two different days and demonstrated significant reproducibility of the HSP platform with overall coefficient of variation averages of 8.82, 8.68, 8.64 and 9.12, for the four QCs, respectively (Figure S2).

The majority of the HSP proteins had similar levels of each HSP protein across the three biofluids based on the overall averages in each of the 8 volunteers biological replicates (Figure 2B). Of note, haptoglobin was higher in plasma than whole blood, however the ratios of haptoglobin in each volunteer relative to the other volunteers followed the same pattern throughout the three biofluids (Figure 3). Subject-specific patterns for other HSP proteins are also seen throughout the biofluids, especially in CRP, VWF, SHBG, IGHA1 as well as other HSP proteins indicating that there were specific proteomic biosignatures in each “healthy/normal” individual that were not affected by the different biofluids. Pearson statistical analysis conducted on all volunteers and all biological replicates also supported the concept that the majority of HSP proteins have similar and reproducible biosignature across biofluids. When capillary blood was directly compared to venous blood there were only three proteins, C1R, SERPINF2 and SERPINC1, that were not statistically similar in the different collection methods, demonstrating that a remote collection device may be used to achieve overall similar results as a traditional phlebotomist blood draw. The variability in the SERPINs may be explained by the activation of blood clotting events in capillary blood during the finger lancet since these proteins are important in controlling proteolytic cascades in coagulation pathways and/or there are less distinct individual differences with these particular proteins compared to more dynamic changes as seen with CRP. As expected, red blood cell proteins HBA and PRDX2, were found in drastically higher concentrations in whole blood compared to plasma which explains the significant difference in the Pearson statistical analysis. In addition, there were additional differences, mostly shared, in venous blood and capillary blood compared to plasma alone. However, it remains to be seen if any of these few differences between biofluids would have an effect on differentiating an individuals’ health within a defined disease group.

Hierarchical clustering heatmaps indicated that each individual volunteer had their own unique biosignature compared to the other volunteers subject specific profiles. Interestingly, three of the volunteers 5, 6, and 7 had elevated HSP proteins that correlated with inflammation. Interestingly, on the same day the volunteers’ samples were collected Volunteer 5 had also had blood collected for the Cedars-Sinai EMBARC study where blood is qualitatively analyzed for IgG against nucleocapsid (N) protein (2.52 where >1.4 is positive) and quantitively analyzed for IgG for the Spike (S) protein (34117.98 AU/mL where ≥50 AU/mL is positive) of Covid-19. The antibody (IgG) results indicate that Volunteer 5 may have had a recent infection with Covid-19. The HSP proteins associated with an elevation of inflammation, CRP, VWF, and HP, and IGHA1 immunoglobin were also elevated in Volunteer 5. Volunteers 6 and 7 also had elevated levels of CRP compared to the other volunteers (Figure 3, 4 and 6A), which also correlated with a history of hypertension and inflammation, respectively. According to the Cleveland Clinic, normal CRP levels are ≤9mg/L and moderate elevation is between 10.0-100.0 mg/L, where >2.0mg/L is at higher risk of heart disease while a hs-CRP test <2.0mg/L is designated lower risk of heart disease. So, although all volunteers CRP levels were below ≤9mg/L, it was interesting to see stratification of the 8 volunteer “healthy/normal” cohort with three individuals potentially correlating with signs of mild inflammation that can be detected with the HSP assay. Overall, the calculated quantitative concentrations of CRP were relatively similar in each biofluid for each volunteer and was within the expected physiological ranges^54,59^ for a normal control cohort, where 5 volunteers CRP levels where <2.0mg/L.

The calibration curves allowed us to calculate the slope for the linear range for each protein based on their HSP SIL peptides. The SIL concentration of 300 fmol was within the linear range of all HSP peptides and therefore we were able to calculate the theoretical quantitation of the endogenous HSP proteins from the overall PAR averages (all 8 volunteers and their biological replicates) in each biofluid (mg/L and nmol/L; Table S7). The PAR for each HSP protein in each individual volunteer can be directly compared to all other individuals throughout any length of time as long as SIL peptides are spiked into each future sample. Interestingly, the calculated concentrations were in the ranges of typical physiological concentrations published in the Plasma Proteome Database http://www.plasmaproteomedatabase.org/)^54^. In the future, intact isotopically labeled proteins could be added to the HSP assay to assess the absolute physiological concentrations of proteins such as with CRP, in order to meet the FDA regulated Tier 1 assay^60^ of physician prescribed blood tests. The dynamic range of HSP proteins in capillary blood (Figure 6C), are all within the lower limits of detection (LLOD).

The unbiased protein discovery assay DIA-MS analysis identified 3,811 unambiguous proteins at 1%FDR (4661 proteins at 5%FDR) in the three biofluids when using the pan-human spectral library^58,60^ compared to the 1,121 proteins identified using a twin plasma spectral library^57^, and 2.21 fold more protein in capillary whole blood and 2.57 fold more protein in venous whole blood, while only a 0.06 fold difference in plasma, demonstrating the significant gain of protein identifications when analyzing whole blood over plasma alone and using a comprehensive spectral library. Interestingly, most HSP proteins (except CRP, SHBG and F10 due to low abundance and likely hidden under interfering peaks from high abundant whole blood proteins, while ITA2B was not detected in any samples) was identified in the whole blood samples demonstrating that the HSP assay could be incorporated into a DIA-hybrid discovery and quantitative assay. More importantly, a larger protein/biomarker multiplex panel could be created with the addition of SILs that represent >122 FDA approved biomarkers, proteins that are already in LDT assays and many of the DIA-MS discovered proteins that have known biological relevance in health and disease, including oxidative stress proteins (e.g. superoxide dismutase, catalase, etc.), longevity (e.g. telomere related proteins), energy metabolism (e.g. creatine kinases, adenylate kinases, etc.), heart and muscle proteins (e.g. Troponin C, myosins, tropomyosins, actin, desmin, FHL2, etc.), oncogene related proteins (e.g. vav, myc, src, etc.), among many other proteins^34^ for a comprehensive biosignature. The biomarker panel could be expanded to include a broad range of low abundant organ specific protein signaling moieties circulating in the bloodstream. These organ derived moieties include myokines (skeletal muscle),^5,61–64^ osteokines (bone),^65,66^ cardiokines (heart),^67^ hepatokines (liver),^68,69^ adipokines (white adipose tissue),^70^ baptokines (brown adipose tissue)^71^ and neurokines (neurons)^72–74^ that may be monitored in health and disease, where intervention through exercise^43^ can stimulate their release, promoting health in other tissues.^75,76^ In addition, the protein activity/function in health and disease can be more specifically monitored through post-translational modifications (PTMs) such as phosphorylation, glycosylation and acetylation as well as monitoring single nucleotide polymorphisms (SNPs) by creating the representative SILs.^77–79^

A growing population of people are taking control of managing their personal health and there has been a rise in companies that are promoting systems to improve consumer access to medical resources and quantifiable diagnostic tests.^80,82–84^ A comprehensive health surveillance panel of proteins would add significant insight into the effectiveness of the treatment, better assess prognosis, and monitor unforeseen side effects. In addition, a comprehensive biomarker panel would help individuals correlate quantitative metrics to the effectiveness of their personalized fitness and nutrition intervention regimes to optimize their health and slow the onset of age-related metabolic diseases^75,76^. The combination of having an easy to use and reliable remote sampling device, to fully utilize the power of mass spectrometry technology advancements to reach an expansive biomarker panel will be critical in the stratification of individual biosignatures for personalized medicine and health.^27,34,80^

## Methods

### Blood and plasma collection

Ethics committee Cedars-Sinai Medical Center (CSMC) IRB No: STUDY00000621 / MOD00004460 CSMC Date Effective: 9/23/2021 gave ethical approval for all human samples used in this study. A total of 8 volunteers signed a consent form to donate venous blood, drawn from a phlebotomist at Cedars-Sinai Medical Center and collected with 10mL K2 EDTA vial. Plasma was isolated from the venous blood by pipetting 500µl into microtainer tubes (lavender, K2 EDTA, tube capacity 250-500µl), inverting 10 times, centrifuging for 3 minutes at 2000xg and pipetting the top layer of plasma into 1.5 ml Eppendorf tubes. In addition, 10µL of venous blood was pipetted onto six Mitra tips for each volunteer. Each volunteer also collected six Mitra device tips (10µl) of their own capillary whole blood from a lancet finger prick. The capillary blood and venous blood was allowed to dry for 30 minutes at room temperature before being stored at 4°C. Mitra Device tips containing capillary whole blood, venous whole blood, and plasma (5.5µl) were added to 2.2mL 96 well plate according to the experiment plate layout (supplemental data). Theoretically, whole blood contains about 55% plasma, therefore we treated each Mitra device containing 10 µl of whole blood as having 5.5µl of plasma (plasma typically contains a protein range concentration between 60-80 µg/µl, therefore PBL treats each volunteers sample to have approximately 70 µg/µl or 5.5µL is about 385µg of plasma for sample processing and direct comparison). Mitra Device tips containing capillary whole blood, venous whole blood, and plasma were added to two 2.2mL 96 well plates (volunteers 1-4 on plate 1 and volunteers 5-8 on plate 2) according to the two experiment plate maps layout (supplemental data). As quality controls commercially purchased pooled plasma (pool4) and pooled plasma from all 8 volunteers was used in triplicate on each 96 well plate. In addition, once capillary blood and venous blood was trypsin digested (see below), an equivalent portion of each volunteers replicates digested peptides were pooled for each biofluid and used as triplicate quality controls.

### Sample Processing

To each Mitra tip 100µL (94.5µL to plasma) of denaturing buffer containing of 67% 100mM ammonium bicarbonate (AmBic) and 33% 2,2,2-Trifluoroethanol (TFE) was added to each of the well. The 96 well plate was sonicated for 15 min at RT, placed on shaker 1000rpm for 30min and centrifuge at 1000 x g for 1 minute. Samples were cysteine reduced by adding 10µL of 55 mM TCEP to the sample(s) and incubating for 30minutes at while shaking at 1000 RPM. Samples were cooled to room temperature for 10 min, centrifuged at 1000 x g for 1 min, before alkylating by adding 10µL of 168mM iodoacetamide (IAM) to the sample(s) and incubating for 20minutes in the dark. Alkylation was quenched by adding 10µL of 65mM dithiothreitol (DTT) to the sample(s) for 15minutes, protected from light. Each sample was diluted with 870µL of 100mM AmBic (final 5% TFE concentration) before adding 10 µL of 1µg/µL trypsin and incubated at 37°C for 16h at 300 rpm. After digestion, samples were cooled to room temperature, centrifuged at 1000 x *g* for 1 minute, before pipetting 144.3µL (equivalent to 50µg of plasma) of each sample into another 96 well plate (according to the plate map), where 855.7µl of H2O was added to bring volume up to 1mL. Stable isotopically labelled (SIL) peptides representing all 60 HSP proteins (see supplemental table) was spiked into each sample solution at 20 µL of 100pmol/mL (2000fmol/50µg of plasma protein). Samples were acidified by adding 20µL of 20% trifluoroacetic acid (TFA) to each sample (∼0.4% TFA) and < pH3 verified with pH indicator strip. A duplicate calibration curve was also added to each MS plate from a dilution series of a HSP SILs spiked into pooled endogenous tryptic digested capillary whole blood from all 8 volunteers six replicate samples before desalting.

The digested and acidified samples were loaded two times 262µL onto a Phenomenex Strata X-PRO desalting plate on top of a 2.2mL 96-well plate, using a pressure manifold set on 2psi. Each well was washed with 1 mL of 5% methanol and eluted into a new 2.2mL 96-well plate with 500 µL of 2% formic acid in 100% methanol (STRATA X-PRO elution buffer). Place the desalting elution plate(s) in the TurboVap (flow rate = 40) until dryness. The dried peptides were solubilized with 100µL of resolubilization buffer (96/4 (v/v) Water/Acetonitrile and 0.2% formic acid) in the desalting elution plate, sealed with foil, sonicated for 5 minutes, shaked at 1000RPM for 10 minutes and then centrifuged 1 minute at 1000 x g. Resolubilized samples were added at 45 µL to two identical mass spectrometry (MS) plates (hard-shell PCR Plates 96 wells and Slit Seal for 96 wells PCR plate) according to MS plate layout (MS plates may also be sealed with foil and stored at −20°C until use). The remaining 10 µL of each sample was pooled into one 1.5 mL tube, mixed, and pipetted into a LC sample vial for LC/MRM-MS retention time correction (RTC).

### HPLC and Mass Spectrometry

LC/MRM-MS was performed on two different systems consisting of ThermoFisher Ultimate 3000 high-flow LC connected to a Sciex 6500+ triple quadrupole mass spectrometer. A System Sensitivity Evaluation was conducted before LC/MRM-MS analysis of samples each using an inhouse PBLQC consisting of a set peptides with variable intensity and retention times. A new batch method in Sciex Analyst containing 2 blanks (0.2% FA) followed by 6 PBLQC (0.2% FA) and 2 blanks with PBLQC 30min MS method to insure sensitivity of LC/MRM-MS before proceeding to RTC, in which all the HSP peptides retention times are corrected from a pool of the remaining tryptic digested samples (the remaining 10µl from each sample not loaded into the duplicate MS plates) and loaded into the Analyst method for scheduled targeted LC/MRM-MS analysis. A sequence of the appropriate number of samples was created in Chromeleon software (Thermo Scientific™ Dionex™Chromeleon™ 7 Chromatography Data System Version 7.2.10) matched to the sample list in Sciex 6500+ Analyst software (Sciex Analyst Instrument Control and Data Processing Software Version 1.7.1) and the targeted HSP peptide and all transitions method with the correct RTC was loaded. Each sample was injected at 15 µL (the equivalent of 7.5µg plasma of capillary blood and venous blood from Mitra tips and 300fmol SIL on column) onto a Waters XBridge Peptide BEH C18 Column, 130Å, 3.5 μm, 2.1 mm × 100 mm. The HPLC gradient duration was 30 min starting at 0 min 5%B 0.500ml/min; 0.800min 5%B to 25 min 30%B at 0.500ml/min; 25.4min 90%B at 0.650ml/min; 25.7min 30%B at 0.650ml/min; 26.1min 90%B at 0.650ml/min; 26.4min 0%B at 0.650ml/min; 26.8min 90%B at 0.650ml/min; 27min 5%B at 0.500ml/min; 30min stop run (Mobile Phase A: 98.8% H^2^O + 0.2% formic acid and Mobile Phase B: 100% Acetonitrile + 0.2% formic acid. LC/MRM-MS data processing and statistical analysis was executed using SCIEX OS (Version 2.1.0.55343) and PBL in-house ProEpic processing platform. All LC/MRM-MS raw files have been deposited to the PASSEL PeptideAtlas repository with the dataset identifier PASS03796.

Data independent acquisitions (DIA) LC-Mass Spec Parameters Analysis was performed on a Thermo UltiMate 3000 LC system coupled to a Thermo Orbitrap Exploris 480. Separation was achieved using the UltiMate 3000 in a trapping configuration, water with 0.1% formic acid as buffer A and acetonitrile with 0.1% formic acid as buffer B. Samples were picked up from the autosampler with a full-loop injection and loaded onto the C18 trap column (10 × 0.3 mm) at 15 µL/min buffer A for 3 minutes. At 3 minutes post-injection, the valve configuration was switched to have the trap column in-line with the analytical column (Phenomenex 15 cm × 300 µm, 100 Å C18 beads), and peptides were separated over a 60-minute gradient at a flow rate of 9.5 µL/min. The concentration of buffer B was increased from 2% to 5% in 2.5 minutes, then to 9% at 6 minutes, 27% at 45 minutes, and 44% at 60 minutes. A nano electrospray source with a NewOmics 8-nozzle emitter (10 µm internal diameter) was used with spray voltage 3800 kV and capillary temperature of 320 °C. MS data on the Orbitrap was collected with a data-independent acquisition (DIA) method with the following parameters: full MS scans were acquired at 60k orbitrap resolution from 400-1000 Da with a normalized AGC target of 300% and maximum injection time of 25 ms. Data-independent fragmentation in MS2 was performed over a mass range of 400-1000 Da using 12 Da windows with an overlap of 1 Da. Peptides were fragmented with a normalized collision energy of 28%. Fragmentation spectra were collected at 15k resolution in the orbitrap from 200-2000 Da with a normalized AGC target of 1000% and maximum injection time of 22 ms. All data was acquired with positive polarity in profile mode. All DIA-MS raw files and search results have been deposited to the ProteomeXchange Consortium via the PRIDE partner repository with the dataset identifier PXD038678.

### Data Analysis and Statistics

PBL in house ProEpic platform was used to analyze raw data, where raw data files information on precursor m/z, product m/z, RT and component name from Analyst are converted (MS Convert) to mzML files that run through SmartPeak software. A custom feature for QC component rules is uploaded specifying 15s width at 50 and a minimum of 6 points across half height was used for all 466 transitions. The MRM report is ranked for quantifier and qualifier peptides reporting peak areas and calculating peak area ratios from endogenous peak area divided by SIL peak area, and calculating %CV.

To assess the linear relationship and magnitude of association for protein intensities across biofluids, Pearson’s Correlation Coefficient (PCC) was calculated using the R programming language (cor.test in stats package). Other statistical measures were evaluated but *PCC* was considered most applicable to the acquired data. Average intensities for each volunteer were compared across three biofluids to produce p value for each comparison in addition to Pearson’s *r*. This calculation was repeated for 57 proteins of interest. The interpretation of the p value indicates statistically significant correlation between the biofluids for a given protein. A calculated p-value < 0.05 equates to a statistically significant positive linear relationship (strong association) between two biofluids. Proteins with significant p values across all biofluid comparisons are indicative of potentially robust biomarkers as they produce more consistent correlation of intensities.

### Reagents and Equipment

In this study the reagents and instruments from Thermo Fisher Scientific: Bond-Breaker TCEP solution (77720B, 0.5 M), Dithiothreitol (DTT, 20291, 7.7mg/tube), Iodoacetamide (IAM, 90034 9.3mg/tube), Methanol (A452-4), formic acid, LC-MS grade (PI28905), microcentrifuge tubes low retention (02-681-320), Template sealing foil (NC9989752), Incubator Thermo Mixer C Eppendorf, Microcentrifuge, Legend Micro 21R, Shaker Digital microplate shaker (model 88880023), Vortex Genie 2 (Sigma Z258423), 96-well plate (2.2 mL, AB0932), MilliporeSigma MQuant pH 0-14 Test Strips, (1095350001), Vial 0.25mL C/T PP 12×32 (Cat# 00278201),11mm orange snap vial cap PTFE/SIL (SUN-SRI, 00250649).

Other reagents and instruments are as indicated in the following: XBridge Peptide BEH C18 Column, 130Å, 3.5 μm, 2.1 mm × 100 mm (186003564 Waters) with a precolumn filter holder (IDEX A-315) and Filter frit, (IDEX A-101X). Strata-X PRO SPE 96-well plate 30 mg/well (8E-S536-TGA, Phenomenex), Hard-shell PCR Plates 96 wells, thin wall Bio Rad #HSP9601 and *Slit Seal for 96 wells PCR plate (190830VO BioChromato*). Centrifuge Allegra X-30R (Beckman Coulter), Human pooled 4 plasma (GW 090220-4; 9/8/2020), Trypsin (LS003741, 500 mg Worthington Biochemical), Mitra Devices (cat# 10101-A Neoteryx), Ammonium Bicarbonate (09830, FW=79.06 Fluka), 2,2,2-Trifluoroethanol (TFE, 75-89-8, Sigma-Aldrich), Trifluoroacetic Acid (TFA, 302031 Sigma-Aldrich), BD Microtainer tubes 500µL (365974 BD), Acetonitrile, HPLC grade (BJAH015-4 VWR International).

## Data Availability

All search results are included in the supplemental excel files. All LC/MRM-MS raw files have been deposited to the PASSEL PeptideAtlas repository with the dataset identifier PASS03796. All DIA-MS raw files and search results have been deposited to the ProteomeXchange Consortium via the PRIDE partner repository with the dataset identifier PXD038678.

http://proteomecentral.proteomexchange.org/cgi/GetDataset?ID=PXD038678

http://www.peptideatlas.org/passel/

## Acknowledgments

We would like to acknowledgments key contributions for the success of this study. We would like to thank Cedars-Sinai for the support of Precision Biomarkers Laboratories (PBL). We would like to thank the phlebotomists at the Human physiology Laboratory and the CORALE study team at Cedars-Sinai for collecting venous blood samples from our 8 volunteers. Shruti Rao coordinated compliance with the IRB protocol, consent forms and volunteer blood draws with Cedars-Sinai phlebotomists as well as provided critical review and comments to the manuscript. We would like to thank Advanced Clinical Biosystems Research Institute (ACBRI) and PBL for previous conceptualization and development of HSP. The original patent “Highly multiplexed and mass spectrometry based methods to measuring 72 human proteins patent application No. 62/448.319 (International publication Number WO, 2018/136825 A1), Inventors Jennifer Van Eyk, Qin Fu, Vidya Venkatraman, and Irina Tchernyshyov. We would also like to thank Qin Fu, Weston Spivia and Esthelle Hoedt for their work in the development and validation of the modified 60 human protein HSP assay in plasma. We would like to thank ACBRI bioinformatics group for the creation, ongoing development and maintaining of ProEpic™ data analysis platform and PBL’s Jonathan Bui’s role and assistance. We would also like to thank Angel Keoseyan for her work with the DIA platform.

## Author contributions

SAW and SMM conceptualized, managed, and planned the execution of the study and are corresponding authors. SAW performed the sample collection, sample processing, LC/MRM-MS experimentation, data analysis, statistical analysis, constructed and finalized all figures, wrote, edited and submitted the final manuscript. SMM provided critical review and comments to the manuscript. AH and AM developed DIA platform, analyzed samples, compiled data and provided critical review and comments to the manuscript. ZLD performed the Pearson correlation coefficient statistical analysis, Pearson result table, Pearson figures and provided critical review and comments to the manuscript. QF is coinventor on the HSP patent used for this study and reviewed the manuscript. JVE is coinventor on the HSP patent, provided key elements for the HSP assay and provided critical review and comments to the manuscript.

## Supplemental Information tables

**Table S1**. HSP 114 peptides and 466 ion transition list for LC/MRM-MS targeted analysis. Delustering potential (DP), dwell time (DT), collision energy (CE), and collision exit potential (CXP).

**Table S2**. Peak area ratio (PAR) and %CV of HSP quantifier peptides from capillary whole blood, venous whole blood and plasma from 8 volunteers.

**Table S3**. Quality control (QC) targeted LC/MRM-MS of HSP peptides on plate 1 and plate 2. Commercially purchased pooled plasma (pool4), pooled capillary whole blood from Mitra tips, pooled venous capillary whole blood from Mitra tips, and pooled plasma from all 8 volunteers biological replicates were analyzed by LC/MRM-MS (PAR) at the beginning, middle and end of both plate 1 and plate 2 on two different instruments on different days.

**Table S4**. Pearson statistical analysis comparing similarity of HSP proteins PAR signal between capillary blood and venous blood, capillary blood and plasma and venous blood and plasma.

**Table S5**. Compilation of volunteer 5’s EMBARC covid-19 antibody levels to the spiked protein and nucleocapsid protein.

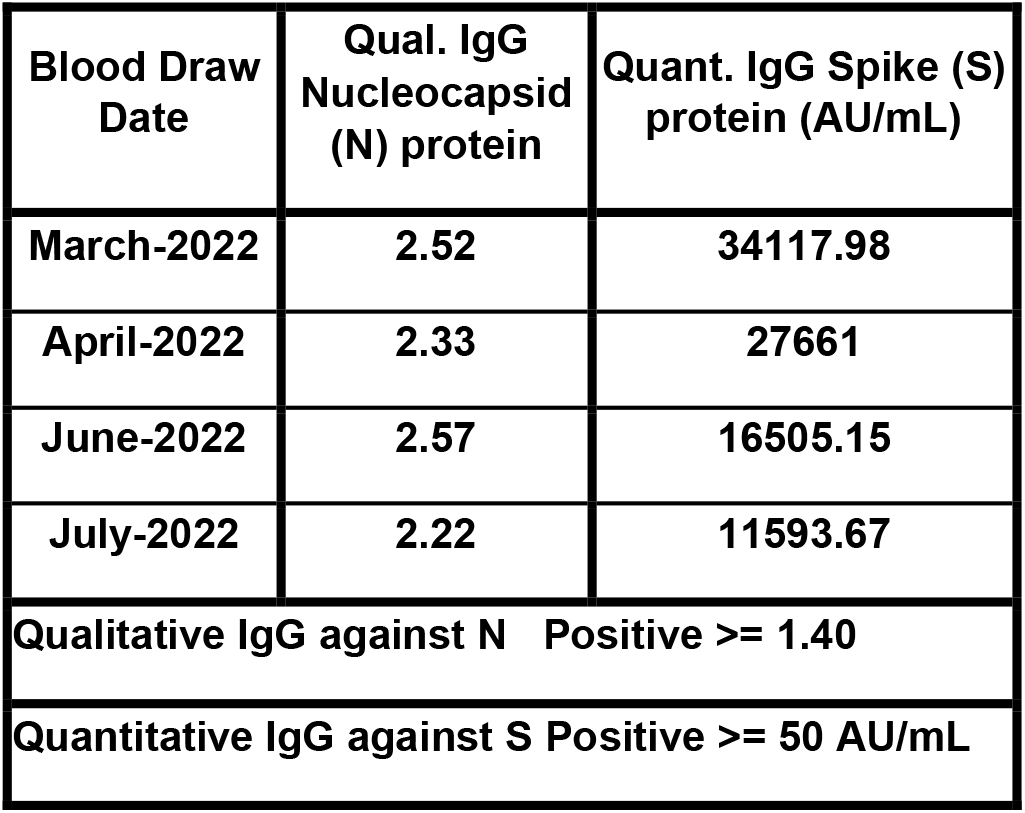

**Table S6**. Average slopes and %CV calculated from calibration curves from plate 1 and plate 2.

**Table S7**. The PAR averages of 8 volunteers and the biological replicates for capillary blood (n=48), venous blood (n=48) and matched plasma (n=24) was used to calculate each HSP protein’s plasma concentration in mg/L and nmol/L. The normal plasma ranges were retrieved from the Plasma Proteome Database to compare the calculated plasma concentrations proteins falling in normal physiological range in plasma are highlighted in green). HSP biomarkers that have been previously approved by the FDA and LDT clinical assays are listed.

**Table S8**. DIA 5% FDR using Twin plasma spectral library.

**Table S9**. DIA data 1% FDR using pan human spectral library. A total of 3881 proteins were identified.

**Table S10**. DIA data 5%FDR using pan human spectral library. A total of 4661 proteins were identified. Capillary blood, venous blood and plasma were ranked based on the sum of peak intensities from the averages of each volunteer for each protein.

## Supplemental Information

**Figure S1.**
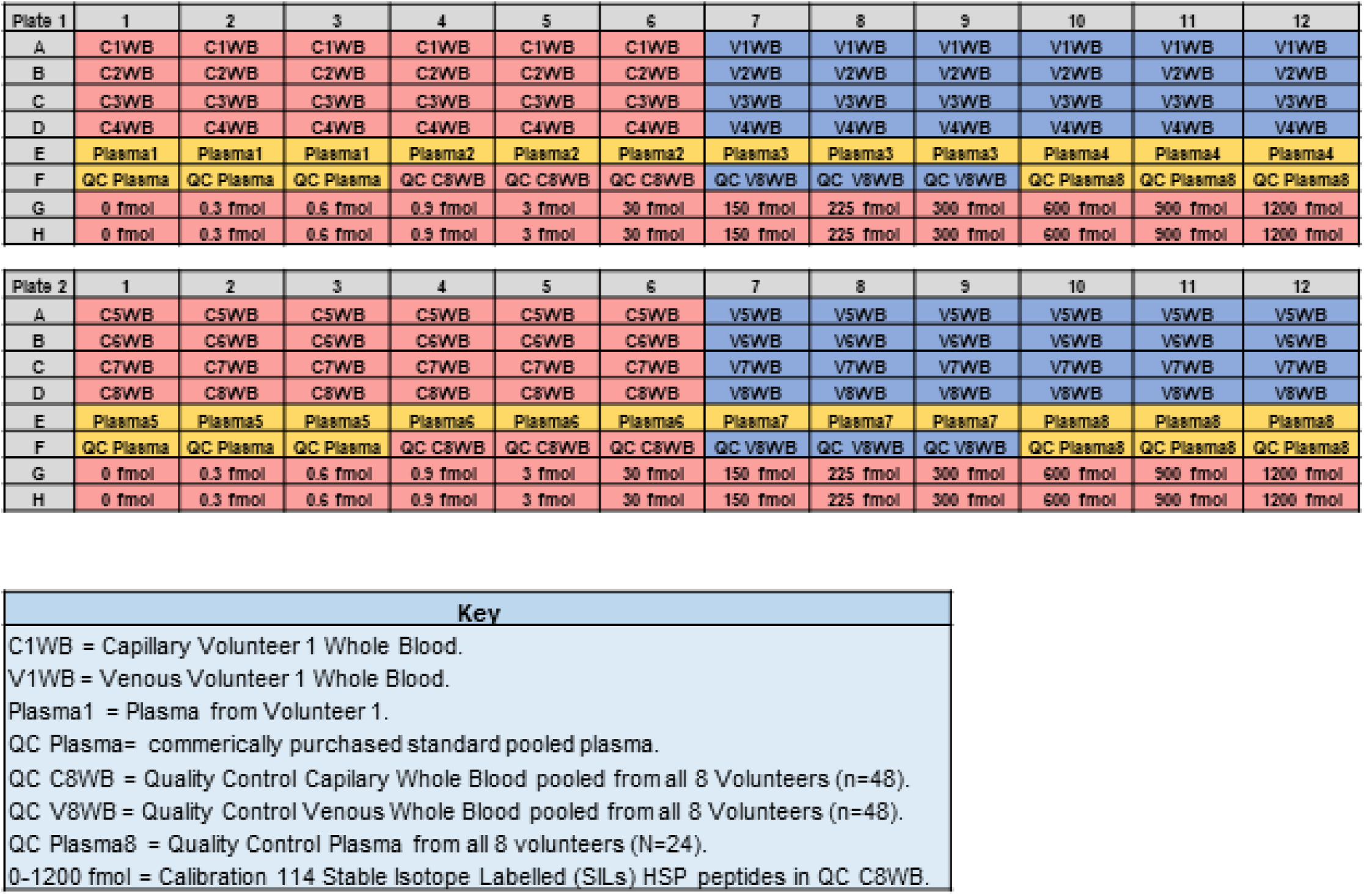
Map of Plate 1 and Plate 2.

**Figure S2.**
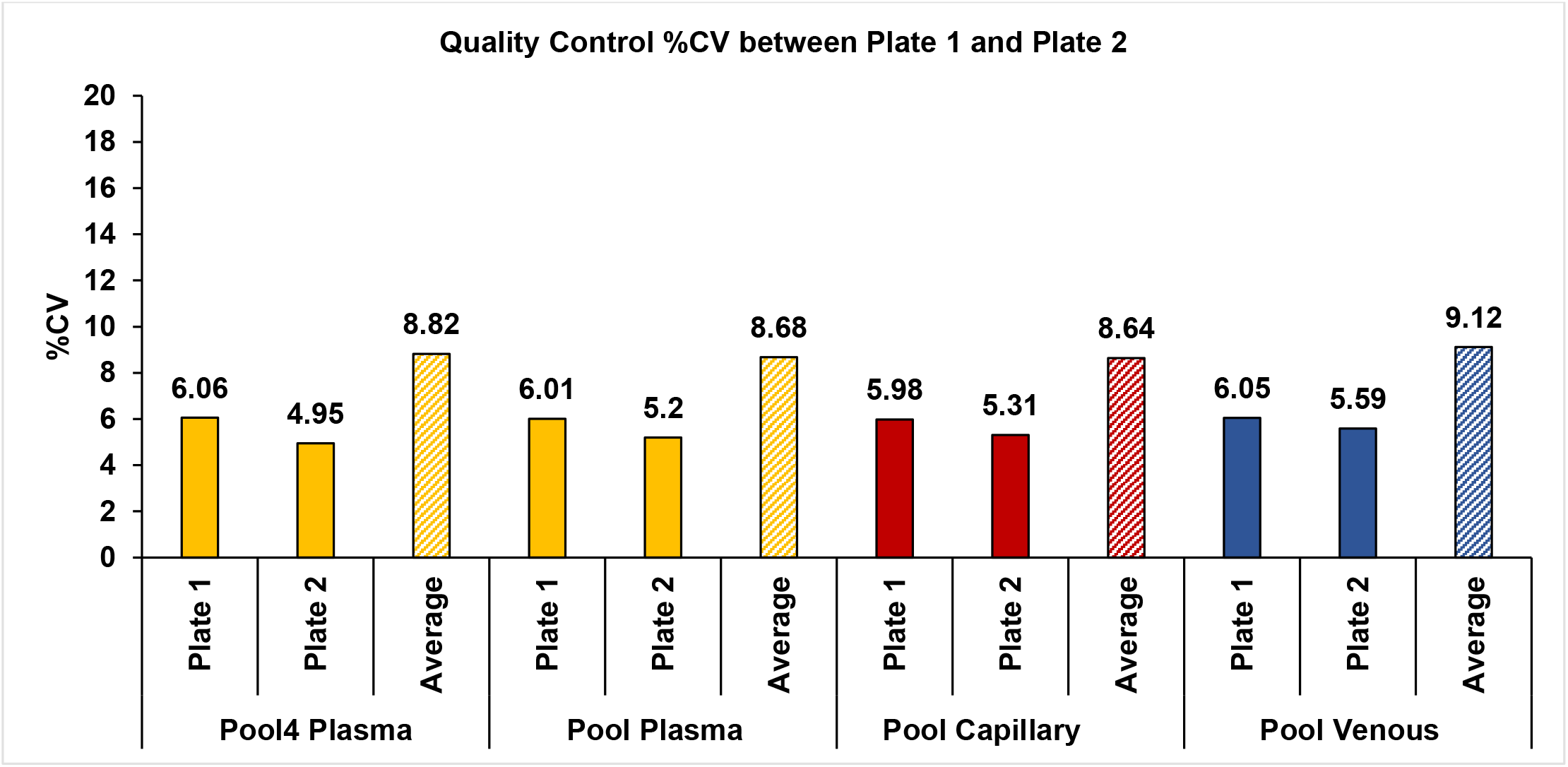
Overall average %CV of HSP proteins in quality control (QC) samples from sample plate 1 and 2. Quality control Below 10% coefficient of variance of Quality Control (QC)samples from two sample plates analyzed on two LC/MRM-MS systems. Quality control (QC) average %CV <10 between plate 1 and plate 2. The QC samples consisted of commercial purchased plasma, pooled volunteer plasma (n=24), pooled volunteer digested capillary blood of all replicates (n=48) and pooled volunteer digested venous blood of all replicates (n=48) analyzed on two different LC/MRM-MS systems in the start, middle and end of both samples plates.

**Figure S3.**
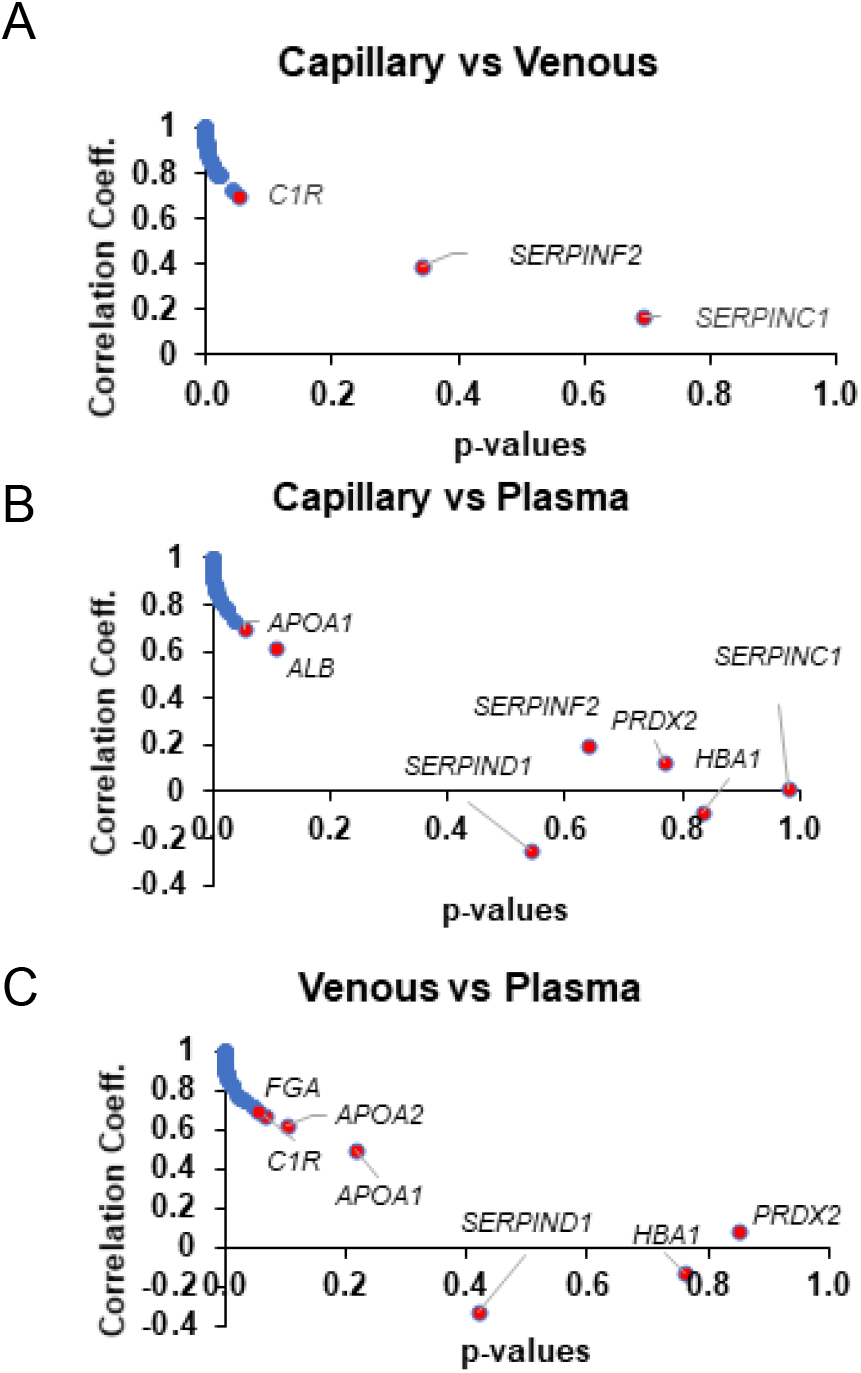
Graphical representation of Pearson statistical analysis results plotting correlation coefficient versus p-values. **(A)** Capillary blood compared to venous blood. B) Capillary blood compared to plasma. **(B)** Venous blood compared to plasma. The majority of HSP proteins p-values are <0.05 (blue circles) indicating significant similarity between compared biofluids. Proteins having p-values >0.05 highlight significance differences (red circles) between the compared biofluids.

**Figure S4.**
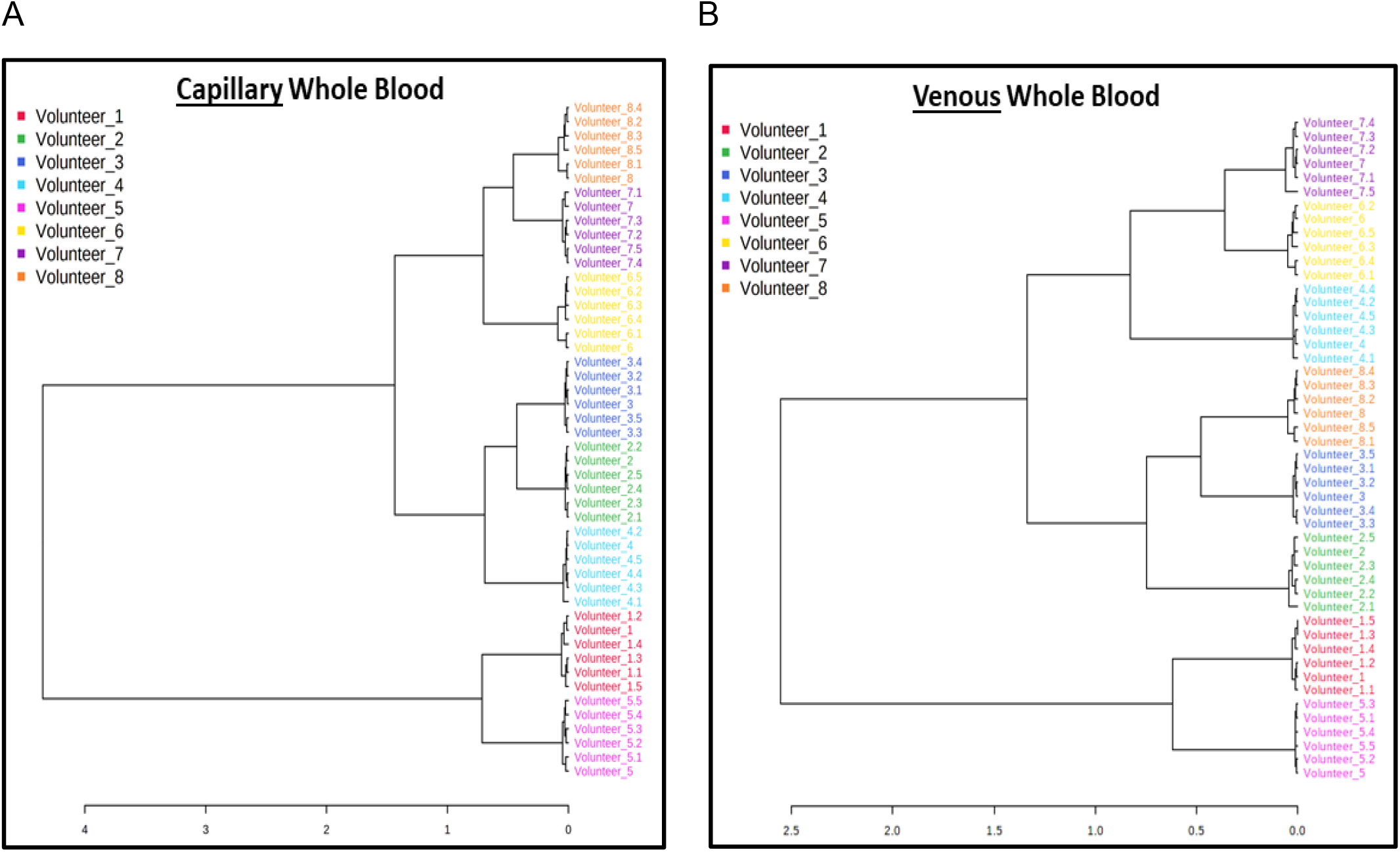
Dendrogram hierarchial clustering of the HSP proteins from each of the 8 volunteers six biological replicate samples. The PAR values were uploaded and analyzed using MetaboAnalyst software for A) capillary blood and B) venous blood samples (distance measure: Spearman; clustering algorithm: Ward).

